# Liver, cardiovascular and metabolic factors as predictors of all-cause mortality in a rural Ugandan Cohort

**DOI:** 10.1101/2025.04.03.25325170

**Authors:** Cori Campbell, Joseph Mugisha, Beatrice Kimono, Elizabeth Waddilove, Ronald Makanga, Tingyan Wang, Florence Nambaziira Muzaale, Janet Seeley, Moffat Nyirenda, Philippa C Matthews, Robert Newton

**Affiliations:** NIHR Oxford Biomedical Research Centre, Oxford, UK; Nuffield Department of Medicine, University of Oxford, Oxford, UK; Medical Research Council/Uganda Virus Research Institute and London School of Hygiene and Tropical Medicine Uganda Research Unit, Entebbe, Uganda; The Francis Crick Institute, London, UK; NIHR Health Informatics Collaborative, Oxford University Hospitals NHS Foundation Trust, Oxford, UK; Social Science Unit, Africa Health Research Institute, KwaZulu-Natal, South Africa. 2Social Science Programme; MRC/UVRI and LSHTM Uganda Research Unit, Entebbe, Uganda; Global Health and Development Department, London School of Hygiene & Tropical Medicine, London, UK; Division of Infection and Immunity, University College London, London, UK; Department of Infectious Diseases, University College London Hospital, London, UK; Department of Health Sciences, University of York, York, United Kingdom

**Keywords:** Liver function tests, liver health, metabolic disease, cardiovascular disease, hypertension, diabetes, MASLD, mortality, HIV, HBV, Uganda, Africa, epidemiology

## Abstract

Markers of liver and metabolic disease have not been well described for many populations in Africa, but could be important to inform individual and public health interventions that reduce morbidity and mortality. We studied longitudinal data from a population in rural Uganda to determine how liver, metabolic and cardiovascular parameters are associated with all-cause mortality. Demographic and laboratory data were collected through the General Population Cohort (GPC) in South-Western Uganda. We summarised cohort characteristics at baseline using descriptive statistics, and used univariable and multivariable Cox proportional hazards models to investigate factors associated with hazards of death. Our dataset includes 7896 individuals, of whom 56% were female and 73% were aged under 45 years. Prevalence of hepatitis B virus (HBV) and human immunodeficiency virus (HIV) was 3% and 7%, respectively, and ALT was above the upper limit of normal in 26%. During the period observed, death was associated with older age (p<0.0001), male sex (aHR 1.56, 95% CI 1.3 to 1.86) and HIV infection (aHR 1.67, 95% CI 1.25 to 2.22). Excess mortality was associated with increase in AST:ALT ratio (aHR 1.17, 95% CI 1.08 to 1.26), a marker of alcoholic hepatitis. A 10mmHg increase in systolic blood pressure was associated with 7% increased hazards of death (aHR 1.07, 95% CI 1.03 to 1.11), and one unit increases in serum HbA1c were associated with 25% increased hazards of death (aHR 1.25, 95% CI 1.13 to 1.38). Under sensitivity analysis restricted to participants with longitudinal follow-up data, HBV infection was associated with death (aHR 5.38, 95% CI 2.01-14.42). Simple interventions to prevent, diagnose and treat hypertension, diabetes, alcohol excess, and HIV and HBV infection could have an important impact on mortality in this setting.

## Introduction

Health surveillance data are urgently needed to better represent populations in the WHO Africa region in order to inform clinical and public health interventions. In this study, we focus on liver parameters, supplemented by cardiovascular and metabolic measurements and biomarkers, to provide a description of a rural East African population to support a better understanding of community health needs and provide an evidence base for intervention.

Hepatic enzymes and biomarkers can be measured in serum to assess liver health and have been applied to measuring the global burden of liver disease^1^. These biomarkers are often collectively referred to as ‘liver function tests’ (LFTs), and include alanine aminotransferase (ALT), aspartate aminotransferase (AST), gamma glutamyl-transferase (GGT), bilirubin (BR) and albumin (Alb). LFT derangement can reflect specific liver pathology, such as viral hepatitis infection, but can also be influenced by systemic perturbations caused by trauma or hypoxia, and be related to extrahepatic or multi-system disorders, including cardiovascular and metabolic disease^2–6^. Updated terminology defines metabolic dysfunction-associated steatotic liver disease (MASLD) based on hepatic steatosis together with cardiovascular and/or metabolic risk factors^7,8^; a growing prevalence of MASLD is increasingly important as a risk factor for mortality worldwide^9^.

While LFTs may be of utility in characterising liver disease and in predicting outcomes, the extent to which different markers are relevant in different population settings is not clearly established^10^. Furthermore, LFT measurement is most commonly undertaken in hospital/secondary care settings, and therefore many studies are not representative of the distribution of LFT values across the whole population. Hypertension, high LDL-cholesterol and elevated haemoglobin A1c (HbA1c) are well-recognised risk factors for cardiovascular mortality, and these parameters need scrutiny in rural African populations to improve targeted monitoring and intervention^11^.

We collected and analysed data from a rural East African setting, collected through the MRC General Population Cohort (GPC), in the South West of Uganda^12^. Our specific objectives were to i) describe the baseline and longitudinal distributions of LFTs, and ii) investigate how LFTs, metabolic and cardiovascular parameters in this setting are associated with all-cause mortality. In the longer term, this investigation can be used to help prioritise screening and interventions for conditions with the greatest impact on public health, supporting resource allocation.

## Methods

### Study design and participant sample

The General Population Cohort (GPC), located in the Ugandan Kyamulibwa sub-county of the Kalungu district (**Figure 1A**), is a prospective population cohort under longitudinal follow up, as previously described^12,13^. The framework was originally established in 1989 to monitor HIV incidence and prevalence in the region, with an annual census to collect demographic, biometric and clinical data. In most waves, all individuals aged ≥13 years planning to reside ≥3 months in a household located within the study region have been eligible for inclusion, with age restrictions relaxed to include younger participants in certain survey rounds. GPC participants have been prospectively followed-up to detect relevant endpoints, and participant data from the survey are linked to data collected at the Medical Research Council (MRC) /Uganda Virus Research Institute (UVRI) clinic via unique individual identifiers. The clinic provides care to participants with acute medical conditions, along with some chronic morbidities including HIV infection and diabetes mellitus.

**Figure 1:**
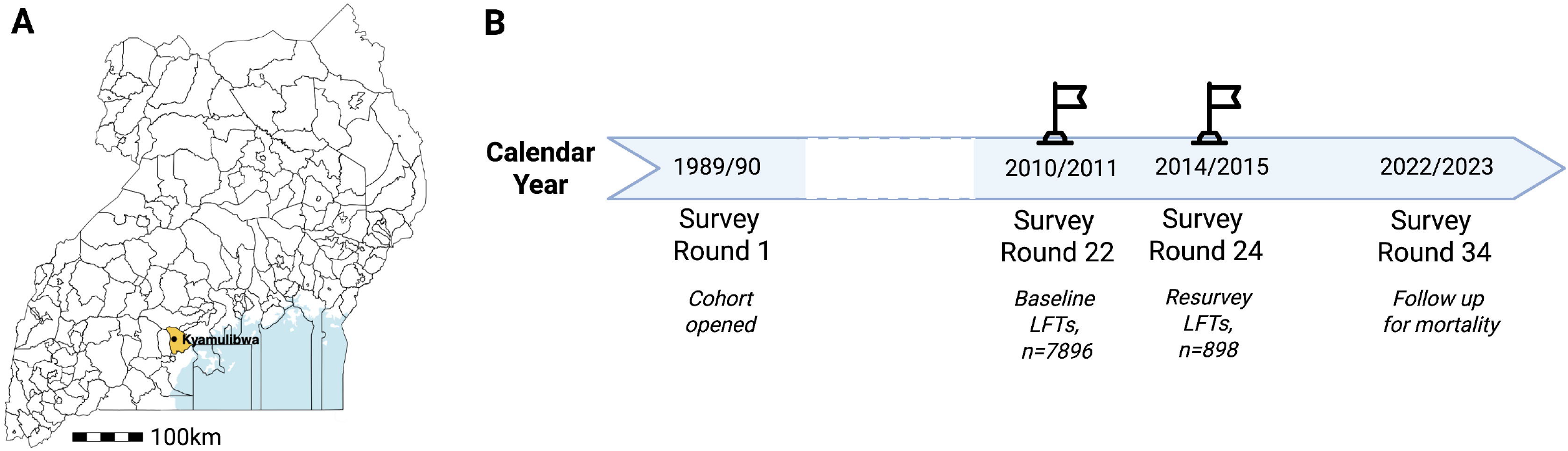
Location and timeline of study of the General Population Cohort in Uganda. [A] Map of Uganda, with county lines marked; Kalungu County is highlighted in yellow and the location of the General Population Cohort in Kyambulibwa is marked. [B] Timepoints of data collection for this study, with flags indicating timepoints at which liver function tests were measured. The cohort was established in 1989, with the study population recruited via annual household census rounds. Follow-up for reporting of deaths is available up to the 2022/23 round for this study.

Data collected in each survey round include demographic factors, biophysical measurements (height, weight, blood pressure and body mass index (BMI, kg/m^2^)). BMI between 18.5 and 24.9 kg/m^2^ was classified as healthy weight, with <18.5 being underweight, and >24.9 overweight. Laboratory analysis of blood samples has been undertaken at intermittent timepoints, subject to availability of funding (not all parameters are measured at every survey round due to cost implications).

For this study, we used LFTs data from survey round 22 (years 2011-12) and LFT resurvey at years 2014-15), which we defined as ‘baseline’ and ‘re-survey’ timepoints respectively (**Figure 1B**). We also report round 22 data for metabolic biomarkers (including HbA1c, high density lipoprotein (HDL), low density lipoprotein (LDL) and triglycerides), and screening for Human Immunodeficiency Virus (HIV) and Hepatitis B Virus (HBV) infection. Children aged 3-12 years were included in round 22, however round 24 was restricted to those aged ≥13 years. Follow-up data for mortality analyses were collected up to years 2022-23.

Socio-demographic data were gathered by interviewers who administered questionnaires to a household head or any adult representative. The World Health Organization (WHO) STEPwise Approach was used to collect cardiovascular risk data^14^. Biophysical measurements (blood pressure, weight, height, waist and hip circumferences) and biochemical analysis (HbA1c, total cholesterol (TC), HDL-C, and triglycerides (TG)) were performed using standardised procedures as previously described^15,16^. The standard approach to blood pressure measurement is described in **S1**.

LFTs (serum AST, ALT, ALP, GGT and BR) were measured in Entebbe, using a Cobas Integra 400 plus machine, with reagents supplied by Roche and protocols according to the manufacturer’s instructions. Methods for testing for Blood Borne Virus Infection (HIV, HBV and HCV) are reported in **S2**.

We used two definitions of LFT derangement to investigate associations with all-cause mortality, (i) applying published ALT thresholds^17^, defined as a serum concentration >19 U/L in females and >30 U/L in males (these thresholds have also employed by international WHO HBV guidelines^18^), and (ii) calculating AST:ALT ratio; where a ratio >2 is an accepted marker of possible alcoholic hepatitis^19,20^. We excluded individuals ≤19 years from ALP analysis, since elevated ALP can be attributable to bone growth in teenagers.

### Follow up and ascertainment of death

The primary outcome for this analysis was death from any cause, which was ascertained from mortality registers, verbal autopsy reports, and reports from community health workers^21^. Mortality registers were updated on a monthly basis over the time period studied.

### Statistical analysis

We summarised cohort characteristics at baseline using descriptive statistics, summarising continuous laboratory parameters with means and standard deviations (SD), or medians and interquartile ranges (IQR), and by division using quintiles. We used univariable and multivariable Cox proportional hazards models to investigate factors associated with hazards of all-cause mortality. The main multivariable model was undertaken in participants surveyed at baseline who had complete information regarding demographic factors, biophysical measurements and markers quantified from blood samples (including LFTs and metabolic biomarkers). Factors were included in multivariable models according to both significance of univariable associations (where *P* ≤0.1) and/or based on biological/clinical relevance and previous literature from both the GPC^12,21^ and investigations undertaken in differing patient populations^3,4,6,22–24^. We calculated univariable Hazard Ratios (HR) and multivariable adjusted Hazards Ratios (aHR), along with 95% confidence intervals (95% CI), as model outputs. Statistical analyses were performed in R (version 4.1.0).

To interrogate the association of BMI with HbA1c in this population, the association between the two variables was visualised, and a nonlinear relationship was fit using a generalised additive model (**S5**).

### Sensitivity analyses

We performed two sensitivity analyses. First, the multivariable Cox proportional hazards model excluded patients who died within three years of baseline LFT measurement, to account for the influence of reverse causality bias on the findings from the main model. Second, we analysed the subset of patients who had LFTs measured at both baseline and resurvey (years 2014-15), to compare the robustness of main-model results against a model accounting for longitudinal changes in LFTs.

### Ethics and reporting

Ethics approval for the GPC was provided by the Science and Ethics Committee of the Uganda Virus Research Institute (GC/127/12/11/06), and the Ugandan National Council for Science and Technology (HS870). All adults provided informed consent for participation including collection of data and samples. Parental/guardian consent for those less than 18 years of age were obtained following Uganda National Council of Science and Technology (UNCST) guidelines. Additional data generation and analysis were approved by the Oxford Tropical Research Ethics Committee (OxTREC; Ref 50-18) for the Uganda Liver Disease Study (ULiDS).

The research has been supported by an active community engagement programme led from the MRC site at Kyambulibwa, which provides health advice and information about research projects, and addresses questions and feedback. Examples of this activity are available in a project report which describes ongoing prospective work on liver disease in this setting^25^. To ensure consistency of reporting, we have included a STROBE (STrengthening the Reporting of OBservational studies in Epidemiology) statement (**S3**).

## Data availability

A data dictionary and data availability can be accessed through the London School of Hygiene and Tropical Medicine Data Compass (https://datacompass.lshtm.ac.uk/id/eprint/4497/; DOI: 10.17037/DATA.00004497)^26^.

## RESULTS

### Baseline characteristics

At baseline, we included data representing 7896 individuals (**Table 1**). There was a slight female excess (4441/7896, 56.2%), and participants aged <45 years accounted for the majority (5722/7896, 72.5%). BMI ranged between 18.5 and 24.9 kg/m^2^ for the majority (4769/7896, 64.8%), representing a healthy range. HBV infection was present in 2.7% (216/7896) and HIV in 7.4% (582/7896). Participants were followed up for a mean 6.4 (SD 3.2) years. At baseline, ALT elevation >ULN was present in 25.6% of this population, and AST:ALT ratio was >2 in 10.9%.

**Table 1.** Baseline characteristics for a subset of individuals included in the Uganda MRC General Population for whom liver function test measurement was performed. Characteristics are reported overall and by survival status after a mean follow up of 6.4 years undertaken between 2010 to 2023. Liver function test resurvey was conducted in a subset of individuals in 2012/2013.

| Characteristic | Total | Died | Survived |
| --- | --- | --- | --- |
| Number of participants | 7896 | 629 (7.97) | 7267 (92.03) |
| Sex |  |  |  |
| Male, n (%) | 3455 (43.8) | 340 (54.1) | 3115 (42.9) |
| Age group, n (%) |  |  |  |
| Age (years), mean (SD) | 34.01 (18.36) | 59.4 (19.5) | 31.8 (16.5) |
| <18 years | 1982 (25.1) | 26 (4.1) | 1956 (26.9) |
| 18-24 years | 1198 (15.2) | 15 (2.4) | 1183 (16.3) |
| 25-34 years | 1379 (17.5) | 48 (7.6) | 1331 (18.3) |
| 35-44 years | 1163 (14.7) | 53 (8.4) | 1110 (15.3) |
| 45-54 years | 936 (11.9) | 75 (11.9) | 861 (11.8) |
| 55-64 years | 578 (7.3) | 107 (17.0) | 471 (6.5) |
| 65-74 years | 414 (5.2) | 152 (24.2) | 262 (3.6) |
| ≥75 years | 246 (3.1) | 153 (24.3) | 93 (1.3) |
| Liver function tests |  |  |  |
| ALT >ULN, n (%) | 2020 (25.6) | 140 (22.3) | 1880 (25.9) |
| AST:ALT ratio, median (IQR) | 1.43 (1.19, 1.71) | 1.64 (1.34, 1.96) | 1.42 (1.18, 1.68) |
| AST:ALT ratio> 2, n (%) | 857 (10.9) | 147 (23.4) | 710 (9.8) |
| ALP, median (IQR) | 92.52<br>(71.23, 144.40) | 90.79<br>(74.85, 116.05) | 92.67<br>(70.82, 150.53) |
| AST >ULN, n (%) | 635 (8.0) | 92 (14.6) | 543 (7.5) |
| GGT, median (IQR) | 1.88 (1.35, 2.79) | 2.58 (1.69, 4.68) | 1.84 (1.33, 2.67) |
| Bilirubin, median (IQR) | 0.76 (0.52, 1.20) | 0.73 (0.49, 1.10) | 0.77 (0.52, 1.21) |
| Albumin, median (IQR) | 41.70<br>(39.25, 43.85) | 38.89<br>(35.00, 41.69) | 41.89<br>(39.57, 44.00) |
| Lipid profile |  |  |  |
| Cholesterol, median (IQR) | 3.41 (2.84, 4.07) | 3.63 (2.94, 4.36) | 3.39 (2.84, 4.05) |
| HDL, median (IQR) | 0.96 (0.74, 1.20) | 1.02 (0.78, 1.32) | 0.95 (0.73, 1.19) |
| LDL, median (IQR) | 1.93 (1.50, 2.43) | 2.00 (1.44, 2.58) | 1.92 (1.50, 2.43) |
| Triglycerides, median (IQR) | 1.05 (0.78, 1.43) | 1.14 (0.85, 1.55) | 1.04 (0.78, 1.41) |
| HbA1c, median (IQR) | 3.33 (2.95, 3.70) | 3.45 (3.04, 3.85) | 3.32 (2.94, 3.69) |
| BMI |  |  |  |
| BMI (kg/m<sup>2</sup>), mean (SD) | 20.66<br>(18.71, 22.90) | 19.74<br>(17.78, 22.22) | 20.70<br>(18.79, 22.95) |
| <18.5 kg/m<sup>2</sup>, n (%) | 1678 (22.8) | 180 (31.9) | 1498 (22.0) |
| 18.5 to 24.9 kg/m<sup>2</sup>, n (%) | 4769 (64.8) | 324 (57.3) | 4445 (65.4) |
| 25 to 29.9 kg/m<sup>2</sup>, n (%) | 718 (9.8) | 42 (7.4) | 676 (9.9) |
| 30 to 34.9 kg/m<sup>2</sup>, n (%) | 147 (2.0) | 13 (2.3) | 134 (2.0) |
| >=35 kg/m<sup>2</sup>, n (%) | 50 (0.7) | 6 (1.1) | 44 (0.6) |
| BP |  |  |  |
| Average systolic BP, median<br>(IQR) | 120<br>(112, 130) | 131<br>(118, 152) | 119<br>(111, 129) |
| Average diastolic BP, median<br>(IQR) | 73<br>(68, 80) | 78<br>(71, 87) | 73<br>(67, 80) |
| HIV status |  |  |  |
| Negative, n (%) | 7314 (92.6) | 564 (89.7) | 6750 (92.9) |
| Positive, n (%) | 582 (7.4) | 65 (10.3) | 517 (7.1) |

| HBV status |  |  |  |
| --- | --- | --- | --- |
| Negative, n (%) | 7680 (97.3) | 611 (97.1) | 7069 (97.3) |
| Positive, n (%) | 216 (2.7) | 18 (2.9) | 198 (2.7) |
| Longitudinal follow-up available |  |  |  |
| Resurveyed, n (%) | 898 (11.4%) | 45 (7.2%) | 853 (11.7%) |

ALT, alanine aminotransferase; ALP, alkaline phosphatase; AST, aminotransferase; IQR, interquartile range; GGT, gamma glutamyl-transferase; HDL, high density lipoprotein; LDL, low density lipoprotein; HbA1c, hemoglobin A1c; BMI, body mass index; BP, blood pressure; HIV, human immunodeficiency virus; HBV hepatitis B virus, ULN, upper limit of normal based on published thresholds<sup>17</sup>.

### Longitudinal LFT trends

LFTs were quantified at re-survey in 898 (11.4%) participants (**Table 2**). Median values of ALT, AST and GGT had fallen compared to baseline, and thus smaller proportions of participants had ALT >ULN^17^ at follow-up (28.7% vs 13.8%, P <0.001). In keeping with the decline in ALT, the median AST:ALT ratio was higher at resurvey, with an increase in the proportion of the population with ratio >2, from 11.2% at baseline to 36.0% at resurvey (P <0.001).

**Table 2.**
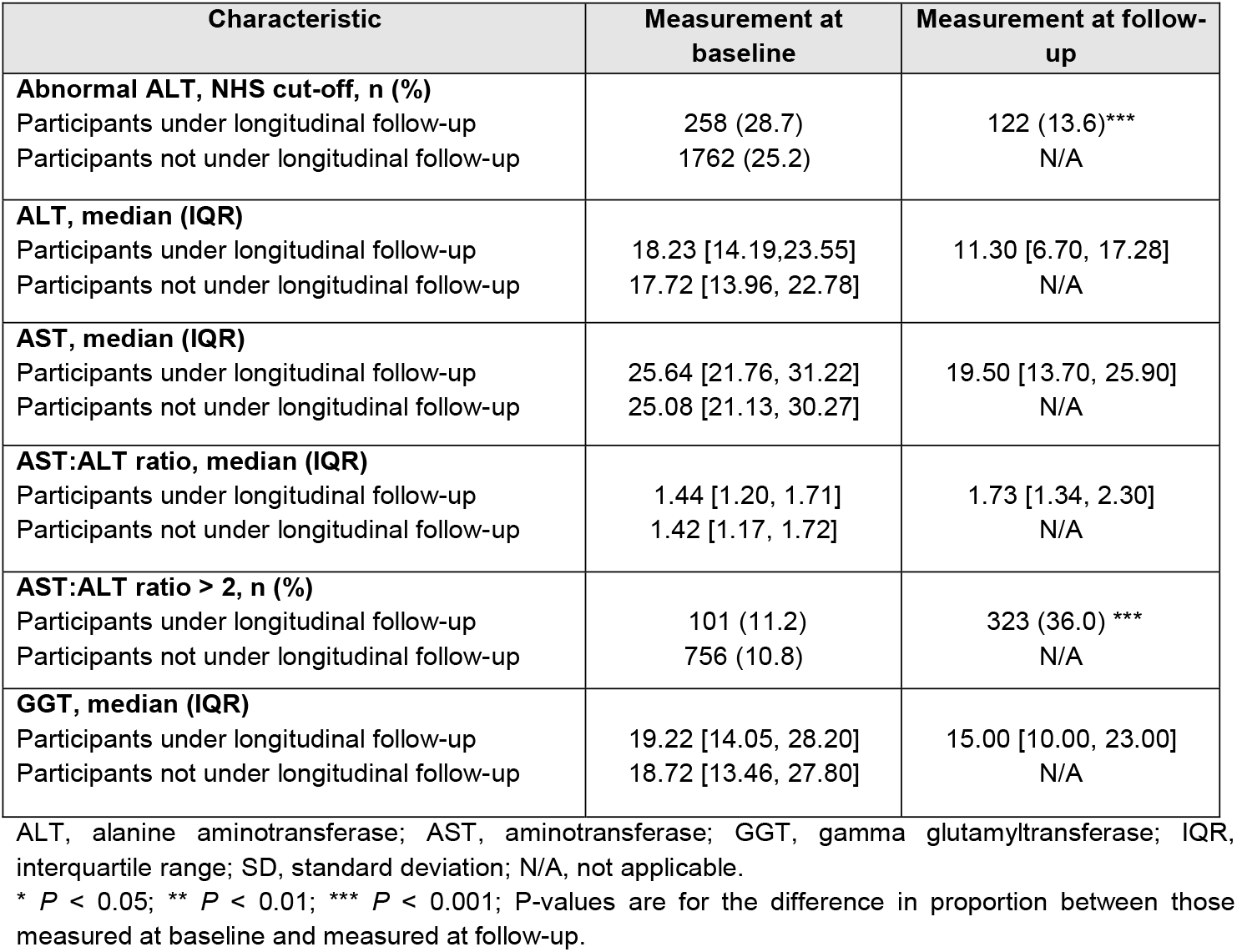
Liver function tests in the Uganda General Population Cohort, divided according to those under longitudinal follow-up and those with baseline measurement only. Baseline data were collected for n = 7896, divided here into those with follow-up measurements (n = 898) and baseline survey data only (n = 6998).

### Risk factors for all-cause mortality

The main complete-case multivariable model included 7341 participants, of whom 563 (7.7%) died throughout follow-up. We identified risk factors for all-cause mortality (**Figure 2**). As expected, mean age was older (59.4 years, SD 19.5) in those who died as compared to those who survived (31.8 years, SD 16.5) (*P* < 0.001). Increased hazards of death during the period of observation were also associated with male sex (aHR 1.56, 95% CI 1.3 to 1.86) and HIV infection (aHR 1.67, 95% CI 1.25 to 2.22).

**Figure 2:**
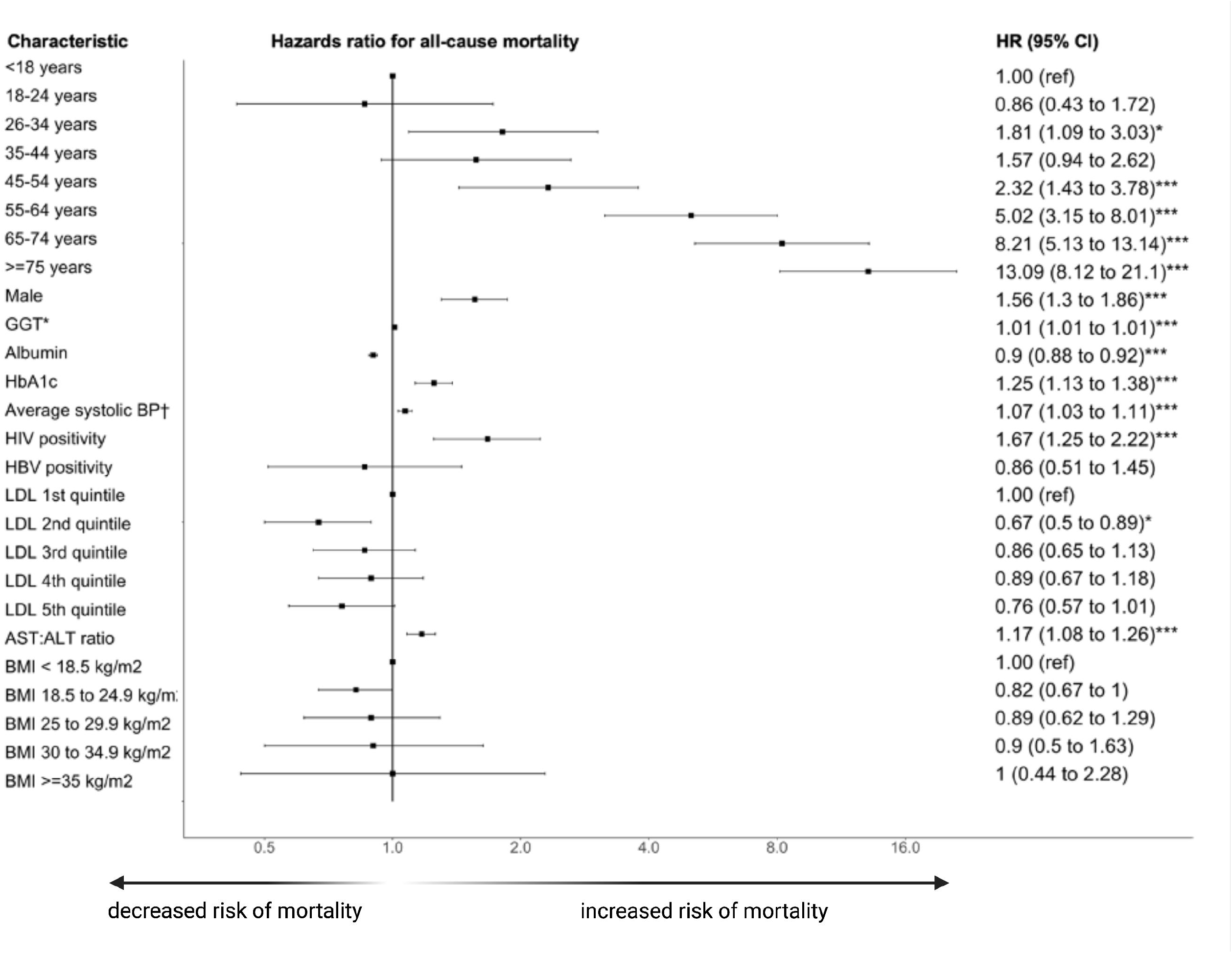
Forest plot for Cox proportional hazards model to identify risk factors for all-cause mortality in the Uganda General Population Cohort study. The plot represents data for 7341 individuals, over a follow-up period of a mean of 6.4 years (SD 3.2 years), among whom there were 563 deaths. This plot includes the estimates for the most parsimonious multivariable model (parameters excluded if univariable associations not significant).

A one unit increase in AST:ALT ratio was associated with 32% increase in death hazards (aHR 1.17, 95% CI 1.08 to 1.26), while 10-unit increases in serum GGT were associated with 1% increased hazards (aHR 1.01, 95% CI 1.01 to 1.01). Both of these markers are associated with alcohol excess. Conversely, hazards were reduced with increased serum albumin, for which a one unit increase predicted a 10% reduction in hazards of death (aHR 0.86, 95% CI 0.85 to 0.87).

On assessment of cardiovascular and metabolic factors, a 10mm Hg increase in systolic blood pressure was associated with 7% increased hazards of death (aHR 1.07, 95% CI 1.03 to 1.11), and one unit increases in serum HbA1c were associated with 25% increased hazards of death (aHR 1.25, 95% CI 1.13 to 1.38). Although HbA1c is associated with BMI (**S5**), there was no statistically significant relationship between BMI and death in this population. Reduced hazards of death were observed in the second LDL quintile (aHR 0.67, 95% CI 0.5 to 0.89) compared to the first quintile, but there were no associations for the other quintile groups.

### Sensitivity analyses

The first sensitivity analysis undertaken was based on data which excluded deaths occurring within the first three years of follow-up (n=5641) (**S4**). In general, the associations observed in the main multivariable model remained unchanged in terms of both direction and magnitude. However, the association of HIV positivity with increasing hazards of death was attenuated towards the null.

The second sensitivity analysis undertaken was restricted to the smaller set of participants with longitudinal LFT measurement (n=871), and utilised a time-updated Cox proportional hazards model to incorporate changes in LFT serum concentrations over time (**S4**). Most associations observed in the main multivariable model remained unchanged in direction. Confidence intervals for all associations were widened due to the smaller number of observations. AST:ALT ratio and HbA1c were no longer significantly associated with mortality in this sensitivity analysis. However, HBV-positive status became significant, with a hazard ratio for mortality of 5.38 (95% CI 2.01-14.42).

## DISCUSSION

### Clinical and public health relevance and rationale

In this young rural Ugandan population, we had the unique opportunity to undertake analysis of longitudinal follow-up in a large cohort, underlining co-endemic HIV and HBV infection, alcohol, and cardiovascular factors as important and modifiable risk factors for mortality. Liver biomarkers that can help to stratify population health include AST:ALT ratio, GGT, and albumin. Hypertension represents a public health concern here, as in many African populations, but recognising that region-specific differences influenced by population genetics, urban/rural location, diet and comorbidity^27,28^. The excess risk of male sex suggests the need for ongoing efforts to target education and interventions towards boys and men, who typically have fewer points of contact with healthcare services than their female counterparts.

Liver, metabolic and cardiovascular related morbidity and mortality are poorly studied in African populations, despite representing a growing public health problem at the intersection of infection and non-communicable disease. In a worldwide study of >1.5million individuals, five modifiable risk factors have been reported to account for ~20% of deaths (BMI, systolic BP, non-HDL cholesterol, smoking and diabetes)^29^. As all of these parameters are also potentially associated with liver disease, and there is growing concern about the global burden of MASLD, we highlight the imperative to collect data representing diverse populations to inform public health strategy. Markers that are associated with mortality and can be measured simply in community settings may help to inform targeted health interventions for education, screening, life-style interventions and treatment.

### Impact of alcohol and metabolic disease on liver health

The combination of alcoholic liver disease (ALD), with metabolic or steatotic liver pathology (‘MetALD’) is increasingly recognised as a global public health concern^30^. The increase in the proportion of our Ugandan cohort with AST:ALT >2 over time, together with a modest but statistically significant relationship between GGT elevation and mortality, is congruent with our previous findings suggesting a high prevalence of alcohol-related liver disease and liver fibrosis in the GPC population^31^. The prevalence and outcomes of ALD and MetALD need further investigation in this setting.

### Chronic viral infection and liver disease

Despite substantial advances in HIV diagnosis, treatment and care, HIV infection in this population was still associated with a significant increase in mortality during the time-course of this study. However, it should be noted that further improvements have continued over time, and updated investigations will be required to determine whether this association has diminished as access to diagnosis and treatment has been strengthened.

HBV has an effect on mortality due mainly to cirrhosis and hepatocellular carcinoma, and is a neglected public health threat in many African populations^32,33^. A significant association between HBV and mortality emerged in our sensitivity analysis (HR 5.38), and our team is undertaking further follow-up of people living with HBV in this population to better understand the characteristics of chronic infection^34^. New WHO guidelines support expanded diagnosis and relaxed treatment eligibility criteria for people living with HBV^18^, and this signal for the relationship with mortality in rural Uganda highlights the pressing case of need for scale-up of HBV testing and treatment. Our data represent an important opportunity for high-profile advocacy for maintaining access to diagnosis, monitoring and treatment in African populations, despite the withdrawal of programmes such as PEPFAR and USAID which have provided life-saving infrastructure, consumables and medications over the past two decades^35^.

### Caveats and limitations

Although the GP aims to represent a cross-section of the whole population, we recognise sources of bias and missingness, as people who are unwell may be less willing or able to enrol in the cohort, and there is migration in and out of the area. At follow-up, LFTs were only measured in a subset of the cohort, due to the cost of running these tests, which affects the power of the study and may account for differences in results between the main and sensitivity analyses. Thresholds for laboratory blood markers and biophysical measurements have not been thoroughly validated in African populations; commonly used thresholds for LFT derangement have been developed in discrete and homogenous populations (more typically in the global North and in high income settings)^17,36^, and therefore may not be sensitive or specific when applied elsewhere^31^.

Our analysis is based on historic data collection, and there is a need for review of new data in order to determine which of the effects we observe remain true over time. We have described associations for which we can offer potential explanations, but factors underlying these associations cannot be definitively determined through this study, and there may be unmeasured confounding that influences our results. We have used all-cause mortality as an endpoint, but further granularity could be added to analysis if causes of death were available; this information was not accessible for this study.

It is important to note that all liver biomarkers are non-specific. Although elevated GGT is often considered a marker of alcohol excess, GGT levels have also been associated with abnormal cardio-metabolic variables in a South African study of people living with HIV^37^, and a change in AST:ALT ratio over time can be driven by improvements in ALT as well as worsening AST. It is therefore important to consider changes in these biomarkers together with other changes in population health over time. Likewise, albumin is a non-specific marker, which when low can be a marker of inadequate nutrition^38^, but is also associated with uncontrolled chronic infection, trauma, malignant or inflammatory disease^39^, and may reflect gut, liver and/or renal disease. Due to the multiple influences that determine serum albumin concentration, there is no single intervention that could be used to target hypoalbuminaemia unless its root causes are more specifically explored.

In this analysis, although HbA1c is associated with increased mortality, population values were low overall. We were underpowered to assess the impact of overweight/obesity due to its low prevalence in the population studied, which may account for the absence of a statistically significant relationship between BMI and mortality in this dataset.

Collection of data on alcohol consumption is important but challenging, as self-reporting is known to be inaccurate, and locally brewed drinks contain varying alcohol concentrations. Locally available alcoholic beverages may also contain other hepatotoxins^40^ which contribute to the acceleration of liver disease.

Assessment of hepatic steatosis requires either a liver biopsy (which is invasive, expensive and carries some clinical risk) or can be quantified using a range of non-invasive tests, including Controlled Attenuation Parameter (CAP) measurement, e.g. using FibroScan^TM^ technology (EchoSens, Paris). The latter is appealing as it is a quick and painless assessment, for which limited operator training is needed, (and subsequent to the timeline of this study) we have introduced this tool for measurement of liver stiffness and steatosis in Uganda^41^ as well as in Kenya and South Africa^42^. However, access to Fibroscan for many settings is currently constrained by the high costs of acquiring and maintaining the hardware.

The associations of increasing age and HIV with increasing hazards of death were attenuated towards the null in the first sensitivity analysis, especially for the older age categories. This likely reflects the influence of reverse causality bias as older participants and those with HIV infection might have had clinically symptomatic and/or advanced disease at recruitment. Only a minority of participants with longitudinal LFT measurement were available for inclusion in sensitivity analyses, which affected our statistical power to detect associations.

## Conclusions

Investing in enhanced assessment of liver health, including viral hepatitis, MASLD, ALD and MetALD is important, as all of these conditions can be tackled through education and public health interventions, with many opportunities for prevention as well as treatment. Male vulnerability may need to be tackled by encouraging and supporting men to attend for screening and health interventions. Further work is needed to determine the extent to which these parameters have continued to influence mortality, in order to inform the delivery of community health programmes.

## Data Availability

A description of the study dataset, as well as access information/instruction for external individuals, is available on the London School of Hygiene and Tropical Medicine Data Compass: https://doi.org/10.17037/DATA.00004497.

## ACKNOWLEDGEMENTS

We are grateful to the study teams and participants in the Uganda General Population Cohort.

## REFERENCES

1. Younossi, Z. M., Wong, G., Anstee, Q. M. & Henry, L. The Global Burden of liver disease. Clin. Gastroenterol. Hepatol. 21, 1978–1991 (2023).

2. Wang, Y., Shi, L., Wang, Y. & Yang, H. An updated meta-analysis of AST and ALT levels and the mortality of COVID-19 patients. Am. J. Emerg. Med. 40, 208–209 (2021).

3. Liu, H. et al. The association between AST/ALT ratio and all-cause and cardiovascular mortality in patients with hypertension. Medicine (Baltimore) 100, e26693 (2021).

4. Wang, T. et al. Longitudinal Analysis of the Utility of Liver Biochemistry as Prognostic Markers in Hospitalized Patients With Corona Virus Disease 2019. Hepatol Commun 5, 1586–1604 (2021).

5. Mayén, A.-L. et al. Hepatic steatosis, metabolic dysfunction and risk of mortality: findings from a multinational prospective cohort study. BMC Med. 22, 221 (2024).

6. Lee, T. H., Kim, W. R., Benson, J. T., Therneau, T. M. & Melton, L. J., 3rd. Serum aminotransferase activity and mortality risk in a United States community. Hepatology 47, 880–887 (2008).

7. Rinella, M. E. et al. A multisociety Delphi consensus statement on new fatty liver disease nomenclature. J. Hepatol. 79, 1542–1556 (2023).

8. Kanwal, F., Neuschwander-Tetri, B. A., Loomba, R. & Rinella, M. E. Metabolic dysfunction-associated steatotic liver disease: Update and impact of new nomenclature on the American Association for the Study of Liver Diseases practice guidance on nonalcoholic fatty liver disease. Hepatology 79, 1212–1219 (2024).

9. Liu, X.-R., Yin, S.-C., Chen, Y.-T. & Lee, M.-H. Metabolic dysfunction-associated steatotic liver disease and its associated health risks. J. Chin. Med. Assoc. 88, 343–351 (2025).

10. Sivakumar, S. et al. Ethnicity-based variations in biological reference interval- A systematic scoping review. Clin. Chim. Acta 578, 120539 (2026).

11. Oni, O. O. et al. Hypertension and its clinical correlates in a rural community in south western Nigeria. West Afr. J. Med. 38, 223–240 (2021).

12. Asiki, G. et al. The general population cohort in rural south-western Uganda: a platform for communicable and non-communicable disease studies. Int. J. Epidemiol. 42, 129–141 (2013).

13. Sekitoleko, I. et al. Identification and characterisation of diabetes in Uganda: protocol for the nested, population-based ‘Diabetes in low-resource Populations’ (DOP) Study. BMJ Open 13, e071747 (2023).

14. The World Health Organization STEPwise approach to non-communicable disease risk factor surveillance. Updated 26 January 2017. https://www.who.int/docs/default-source/ncds/ncd-surveillance/steps/steps-manual.pdf.

15. Asiki, G. et al. Prevalence of dyslipidaemia and associated risk factors in a rural population in South-Western Uganda: a community based survey. PLoS One 10, e0126166 (2015).

16. Murphy, G. A. et al. Sociodemographic distribution of non-communicable disease risk factors in rural Uganda: a cross-sectional study. Int. J. Epidemiol. 42, 1740–1753 (2013).

17. Newsome, P. N. et al. Guidelines on the management of abnormal liver blood tests. Gut 67, 6–19 (2018).

18. Guidelines for the prevention, diagnosis, care and treatment for people with chronic hepatitis B infection. Geneva: World Health Organization; 2024. Licence: CC BYNC-SA 3.0. https://www.who.int/publications/i/item/9789240090903.

19. Torruellas, C., French, S. W. & Medici, V. Diagnosis of alcoholic liver disease. World J. Gastroenterol. 20, 11684–11699 (2014).

20. Botros, M. & Sikaris, K. A. The de ritis ratio: the test of time. Clin. Biochem. Rev. 34, 117–130 (2013).

21. Kalyesubula, R. et al. Association of impaired kidney function with mortality in rural Uganda: results of a general population cohort study. BMJ Open 12, e051267 (2022).

22. Kazemi-Shirazi, L. et al. Gamma glutamyltransferase and long-term survival: is it just the liver? Clin. Chem. 53, 940–946 (2007).

23. Peltz-Sinvani, N. et al. Low ALT levels independently associated with 22-year all-cause mortality among coronary heart disease patients. J. Gen. Intern. Med. 31, 209–214 (2016).

24. Hernaez, R. et al. Elevated ALT and GGT predict all-cause mortality and hepatocellular carcinoma in Taiwanese male: a case-cohort study. Hepatol. Int. 7, 1040–1049 (2013).

25. Kimono, B. et al. Uganda Liver Disease Study (ULiDS) - Project Summary 2023-2024. (figshare, 2024). doi:10.6084/M9.FIGSHARE.25377379.

26. Newton, R. et al. Uganda Liver Disease study – Dataset for analysis of liver, metabolic and cardiovascular parameters associated with mortality in the General Population Cohort. London School of Hygiene & Tropical Medicine 10.17037/DATA.00004497 (2025).

27. Ayalew, T. L., Wale, B. G. & Zewudie, B. T. A systemic review and meta-analysis on the prevalence and associated factors of hypertension among adult clients in Ethiopia. Afr. Health Sci. 23, 296–314 (2023).

28. Ranzani, O. T. et al. Urban-rural differences in hypertension prevalence in low-income and middle-income countries, 1990-2020: A systematic review and meta-analysis. PLoS Med. 19, e1004079 (2022).

29. Global Cardiovascular Risk Consortium et al. Global effect of modifiable risk factors on cardiovascular disease and mortality. N. Engl. J. Med. 389, 1273–1285 (2023).

30. Gratacós-Ginès, J., Ariño, S., Sancho-Bru, P., Bataller, R. & Pose, E. MetALD: Clinical aspects, pathophysiology and treatment. JHEP Rep. 7, 101250 (2025).

31. O’Hara, G. et al. Liver function tests and fibrosis scores in a rural population in Africa: a cross-sectional study to estimate the burden of disease and associated risk factors. BMJ Open 10, e032890 (2020).

32. O’Hara, G. A. et al. Hepatitis B virus infection as a neglected tropical disease. PLoS Negl. Trop. Dis. 11, e0005842 (2017).

33. Delphin, M. et al. Under-representation of the WHO African region in clinical trials of interventions against hepatitis B virus infection. Lancet Gastroenterol. Hepatol. 9, 383–392 (2024).

34. Lumley, S. F. et al. Eligibility for hepatitis B virus (HBV) treatment and prevalence of drug resistance in a Ugandan population cohort. medRxiv (2025) doi:10.1101/2025.04.06.25325341.

35. Wang, S. et al. Pathogens don’t respect politicians: US federal disruption poses a new threat to global public health. Lancet Gastroenterol. Hepatol. 10, 291–292 (2025).

36. European Association for the Study of the Liver. EASL Clinical Practice Guidelines on non-invasive tests for evaluation of liver disease severity and prognosis - 2021 update. J. Hepatol. 75, 659–689 (2021).

37. Nguyen, K. A., Peer, N. & Kengne, A. P. Associations of gamma-glutamyl transferase with cardio-metabolic diseases in people living with HIV infection in South Africa. PLoS One 16, e0246131 (2021).

38. Soeters, P. B., Wolfe, R. R. & Shenkin, A. Hypoalbuminemia: Pathogenesis and clinical significance. JPEN J. Parenter. Enteral Nutr. 43, 181–193 (2019).

39. Buitendag, J. et al. Serum albumin nadir as marker of inflammatory response in abdominal trauma. S. Afr. J. Surg. 62, 40–44 (2024).

40. Otim, O., Juma, T. & Otunnu, O. Assessing the health risks of consuming ‘sachet’ alcohol in Acoli, Uganda. PLoS One 14, e0212938 (2019).

41. Lumley, S. et al. Liver elastography scores in a large population cohort in rural Uganda: Uganda Liver Disease Study (ULiDS). (2024) doi:10.6084/M9.FIGSHARE.25377049.

42. Anderson, M. et al. ELASTOGRAPHY AND CAP ASSESSMENT FOR LIVER DISEASE: Training events and feedback representing clinical research programmes in South Africa, Uganda and Kenya. Preprint at 10.6084/M9.FIGSHARE.27941958 (2024).

