## Supplementary material for "Liver, cardiovascular and metabolic factors as predictors of all-cause mortality in a rural Ugandan Cohort"

### CONTENTS OF SUPPLEMENT


**S1:** Standardised approach to blood pressure measurement P2

**S2:** Standardised approach to testing for blood borne virus infection P2

**S3:** STROBE statement P3

**S4:** Results of sensitivity analysis P7

**S5.** Relationship between body mass index and HbA1c P9

**S1: Standardised approach to blood pressure measurement**

Blood pressure was measured on the right arm using appropriate cuff sizes (regular arm cuff size if arm circumference was 24-32cm, large arm cuff if arm circumference was 33-41cm, thigh cuff if over 41cm and if under 24cm paediatric cuff size was used) in a sitting position, three times within resting intervals of 5 minutes, using a digital sphygmomanometer (the Omron M4-I). The mean of the second and third reading was taken for analysis. Body weight was measured using the Seca 761 mechanical scales and body height was measured using a portable Leicester stadiometer to the nearest 1 kg and 0.1 cm respectively, with participants wearing light clothing and no shoes.

**S2: Standardised approach to testing for blood borne virus infection**

HIV testing was done using an algorithm recommended by the Uganda Ministry of Health, starting with screening with a locally approved rapid diagnostic test (RDT). If the test result was negative, the participant was considered to be HIV negative. If the test result was positive, the sample was retested with the rapid test HIV-1 or HIV-2 Stat-Pak. If both tests resulted in a positive result, the participant was diagnosed as living with HIV infection. If the tests gave discordant results, the sample was further evaluated with the rapid test Uni-Gold Recombinant HIV-1/2. For those samples assessed by all three tests, two positive test results were interpreted as positive, and two negative results were considered negative. HBV surface antigen (HBsAg) testing was conducted using Cobas HBsAg II (2011–08 V.10), and those who tested positive were invited for further serologic testing. HCV was tested using a combination of immunoassays followed by PCR, as previously described*.

* O'Hara G, et al. Liver function tests and fibrosis scores in a rural population in Africa: a cross-sectional study to estimate the burden of disease and associated risk factors. BMJ Open. 2020 ;10(3):e032890. Available from: https://bmjopen.bmj.com/content/10/3/e032890

**S3. STROBE Statement.**

| Section in Paper | Item No. | Recommendation | Notes |
|---|---|---|---|
| **Title and abstract** | 1 | (*a*) Indicate the study's design with a commonly used term in the title or the abstract | Cohort study indicated in title |
| | | (*b*) Provide in the abstract an informative and balanced summary of what was done and what was found | Abstract presented to include background, methods, key findings and rationale. |
| Introduction | | | |
| **Background/rationale** | 2 | Explain the scientific background and rationale for the investigation being reported | Introduction explains case of need. |
| **Objectives** | 3 | State specific objectives, including any prespecified hypotheses | Specific aims stated at the end of introduction section |
| Methods | | | |
| **Study design** | 4 | Present key elements of study design early in the paper | Study design presented in methods with figure 1 to show timeline. |
| **Setting** | 5 | Describe the setting, locations, and relevant dates, including periods of recruitment, exposure, follow-up, and data collection | Details included in methods. |
| **Participants** | 6 | (*a*) Give the eligibility criteria, and the sources and methods of selection of participants. Describe methods of follow-up | Details included in methods and through reference to other papers. |
| | | (*b*) For matched studies, give matching criteria and number of exposed and unexposed | N/A |
| **Variables** | 7 | Clearly define all outcomes, exposures, predictors, potential confounders, and effect modifiers. Give diagnostic criteria, if applicable | Relevant markers described in methods section. Ascertainment of primary outcome (mortality) described in methods. |
| **Data sources / measurement** | 8* | For each variable of interest, give sources of data and details of methods of assessment (measurement). Describe comparability of assessment methods if there is more than one group | Measurements, variables and reference ranges (where relevant) stated in methods. |

| | | | |
|---|---|---|---|
| **Bias** | 9 | Describe any efforts to address potential sources of bias | Bias and missingness addressed in the discussion section. |
| **Study size** | 10 | Explain how the study size was arrived at | N/A – we used full data from a community population cohort (described in methods). |
| **Quantitative variables** | 11 | Explain how quantitative variables were handled in the analyses. If applicable, describe which groupings were chosen and why | Handling of quantitative data is described in methods. |
| **Statistical methods** | 12 | (*a*) Describe all statistical methods, including those used to control for confounding | Statistical approaches presented in methods. |
| | | (*b*) Describe any methods used to examine subgroups and interactions | Statistical approaches presented in methods. No subgroups or interactions were examined, but sensitivity analysis was undertaken to test robustness of the main analysis to reverse causality bias. |
| | | (*c*) Explain how missing data were addressed | Statistical approaches presented in methods. Sensitivity analysis undertaken in patients who were resurveyed after baseline measurement. |
| | | (*d*) If applicable, explain how loss to follow-up was addressed | Statistical approaches presented in methods. Verbal autopsies are undertaken to ensure completeness of death records. |
| | | (*e*) Describe any sensitivity analyses | Sensitivity analyses described. |
| Results | | | |
| **Participants** | 13* | (a) Report numbers of individuals at each stage of study—eg numbers potentially eligible, examined for eligibility, confirmed eligible, included in the study, completing follow-up, and analysed | Numbers are described and presented in results, tables and figure 1. |
| | | (b) Give reasons for non-participation at each stage | Description of loss to follow-up is presented. |
| | | (c) Consider use of a flow diagram | Flow diagram (timeline) is included. |

| | | | |
|---|---|---|---|
| **Descriptive data** | 14* | (a) Give characteristics of study participants (eg demographic, clinical, social) and information on exposures and potential confounders | Descriptive data are presented in Table 1. |
| | | (b) Indicate number of participants with missing data for each variable of interest | Number of participants under follow up presented in Table 2. |
| | | (c) Summarise follow-up time (eg, average and total amount) | Follow-up time presented in results and in Figure 1. |
| **Outcome data** | 15* | Report numbers of outcome events or summary measures over time | Outcome events (mortality) presented in results section. |
| **Main results** | 16 | (*a*) Give unadjusted estimates and, if applicable, confounder-adjusted estimates and their precision (eg, 95% confidence interval). Make clear which confounders were adjusted for and why they were included | Presented in tables. |
| | | (*b*) Report category boundaries when continuous variables were categorized | Presented in methods. |
| | | (*c*) If relevant, consider translating estimates of relative risk into absolute risk for a meaningful time period | N/A |
| **Other analyses** | 17 | Report other analyses done—eg analyses of subgroups and interactions, and sensitivity analyses | Sensitivity analyses conducted and presented in supplementary table. |
| Discussion | | | |
| **Key results** | 18 | Summarise key results with reference to study objectives | Presented in results and discussion. |
| **Limitations** | 19 | Discuss limitations of the study, taking into account sources of potential bias or imprecision. Discuss both direction and magnitude of any potential bias | Presented in discussion. |
| **Interpretation** | 20 | Give a cautious overall interpretation of results considering objectives, limitations, multiplicity of analyses, results from similar studies, and other relevant evidence | Presented in discussion. |
| **Generalisability** | 21 | Discuss the generalisability (external validity) of the study results | Presented in discussion. |

| Other information | | | |
|---|---|---|---|
| **Funding** | 22 | Give the source of funding and the role of the funders for the present study and, if applicable, for the original study on which the present article is based | Funding information is presented at the end of the article. |

**S4. Cox proportional hazards model for mortality based on univariable associations, complete case main model (n = 7341, deaths = 563) and sensitivity analyses.** Sensitivity analysis I: excluding deaths occurring in the first 3 years of follow-up (n = 5641, deaths = 427). Sensitivity analysis II: restricted to participants with longitudinal LFT measurement (n = 871, deaths = 44).

| Characteristic | Univariable HR (95% CI) | Multivariable HR (95% CI) | Multivariable HR sensitivity analysis I (95% CI) | Multivariable HR sensitivity analysis II (95% CI) |
|---|---|---|---|---|
| Age group, n (%) | | | | |
| <18 years | 1.00 (ref) | 1.00 (ref) | 1.00 (ref) | 1.00 (ref) |
| 18-24 years | 0.89 (0.47 to 1.69) | 0.86 (0.43 to 1.72) | 0.6 (0.26 to 1.36) | 2.17 (0.13 to 35.18) |
| 25-34 years | 1.72 (1.07 to 2.78)* | 1.81 (1.09 to 3.03)* | 1.2 (0.68 to 2.14) | 5 (0.62 to 40.37) |
| 35-44 years | 1.96 (1.23 to 3.14)** | 1.57 (0.94 to 2.62) | 1.18 (0.67 to 2.06) | 2.17 (0.24 to 19.84) |
| 45-54 years | 3.28 (2.1 to 5.14)*** | 2.32 (1.43 to 3.78)*** | 1.85 (1.09 to 3.14)* | 7.56 (0.89 to 64.15) |
| 55-64 years | 7.1 (4.62 to 10.93)*** | 5.02 (3.15 to 8.01)*** | 3.9 (2.35 to 6.48)*** | 8.73 (1.05 to 72.87) |
| 65-74 years | 14.87 (9.79 to 22.59)*** | 8.21 (5.13 to 13.14)*** | 6.16 (3.68 to 10.31)*** | 7.03 (0.82 to 60.01) |
| ≥75 years | 25.04 (16.4 to 38.23)*** | 13.09 (8.12 to 21.1)*** | 9.16 (5.41 to 15.5)*** | 31.77 (3.83 to 263.64)*** |
| Sex, n (%) | | | | |
| Female | 1.00 (ref) | 1.00 (ref) | 1.00 (ref) | 1.00 (ref) |
| Male | 1.42 (1.22 to 1.67)*** | 1.56 (1.3 to 1.86)*** | 1.49 (1.22 to 1.83)*** | 2.12 (1.01 to 4.45)* |
| LFTs, cardiovascular and metabolic markers | | | | |
| GGT* | 1.01 (1.01 to 1.02)*** | 1.01 (1.01 to 1.01)*** | 1.01 (1.00 to 1.01)* | 1.02 (0.95 to 1.08) |
| AST:ALT ratio | 1.32 (1.26 to 1.39)*** | 1.17 (1.08 to 1.26)*** | 1.23 (1.11 to 1.36)*** | 0.87 (0.58 to 1.3) |
| Albumin | 0.86 (0.85 to 0.87)*** | 0.9 (0.88 to 0.92)*** | 0.92 (0.9 to 0.94)*** | 0.91 (0.86 to 0.97)*** |
| HbA1c | 1.25 (1.16 to 1.35)*** | 1.25 (1.13 to 1.38)*** | 1.23 (1.07 to 1.41)*** | 0.89 (0.57 to 1.4) |
| Average systolic BP † | 1.25 (1.21 to 1.29)*** | 1.07 (1.03 to 1.11)*** | 1.08 (1.03 to 1.13)*** | 1.21 (1.07 to 1.38)*** |
| LDL quintiles | | | | |
| First | 1.00 (ref) | 1.00 (ref) | 1.00 (ref) | 1.00 (ref) |
| Second | 0.59 (0.46 to 0.77)*** | 0.67 (0.5 to 0.89)* | 0.69 (0.49 to 0.96)* | 0.68 (0.27 to 1.71) |

| | | | | |
|---|---|---|---|---|
| Third | 0.74 (0.58 to 0.95)* | 0.86 (0.65 to 1.13) | 0.93 (0.67 to 1.28) | 0.65 (0.25 to 1.71) |
| Fourth | 0.76 (0.6 to 0.97)* | 0.89 (0.67 to 1.18) | 0.93 (0.67 to 1.28) | 0.77 (0.32 to 1.84) |
| Fifth | 0.81 (0.64 to 1.01) | 0.76 (0.57 to 1.01) | 0.72 (0.52 to 1) | 0.42 (0.13 to 1.36) |
| HIV status | | | | |
| HIV positivity | 1.41 (1.09 to 1.82)** | 1.67 (1.25 to 2.22)*** | 1.31 (0.91 to 1.88) | 1.89 (0.65 to 5.44) |
| HBV positivity | 0.95 (0.59 to 1.52) | 0.86 (0.51 to 1.45) | 0.84 (0.47 to 1.51) | 5.38 (2.01 to 14.42)*** |
| BMI, n (%) | | | | |
| Underweight (<18.5 kg/m$^2$) | 1.00 (ref) | 1.00 (ref) | 1.00 (ref) | 1.00 (ref) |
| Normal weight (18.5-24.9 kg/m$^2$) | 0.63 (0.52 to 0.75)*** | 0.82 (0.67 to 1) | 0.94 (0.75 to 1.19) | 0.57 (0.3 to 1.09) |
| Overweight (25.0-29.9 kg/m$^2$) | 0.56 (0.4 to 0.79)*** | 0.89 (0.62 to 1.29) | 0.96 (0.63 to 1.47) | 0.25 (0.03 to 2.01) |
| Obesity class (30.0 to 34.9 kg/m$^2$) | 0.7 (0.4 to 1.23) | 0.9 (0.5 to 1.63) | 0.92 (0.47 to 1.82) | 1.27 (0.18 to 8.93) |
| Obesity classes II and III (≥35 kg/m$^2$) | 1.01 (0.44 to 2.27) | 1 (0.44 to 2.28) | 1.42 (0.62 to 3.3) | 2.45 (0.21 to 29.07) |

HR, hazards ratio; CI, confidence interval; GGT, gamma glutamyltransferase; HbA1c, hemoglobin A1c; BP, blood pressure; HIV, human immunodeficiency virus; HBV hepatitis B virus; LDL, low density lipoprotein; ALT, alanine aminotransferase; AST, aminotransferase; BMI, body mass index;.

* Hazards ratio per 10 unit increase in serum GGT.

† Hazards ratio per 10 unit increase in systolic blood pressure.

* $P < 0.05$

** $P < 0.01$

*** $P < 0.001$

**S5. Relationship between body mass index and HbA1c in the Uganda GPC.** As anticipated, as BMI increases, so does HbA1c, but HbA1c values in this population are low (confidence intervals do not span the clinically important thresholds of 5.7% which represents a cut-off for pre-diabetes and 6.5% which indicates diabetes).

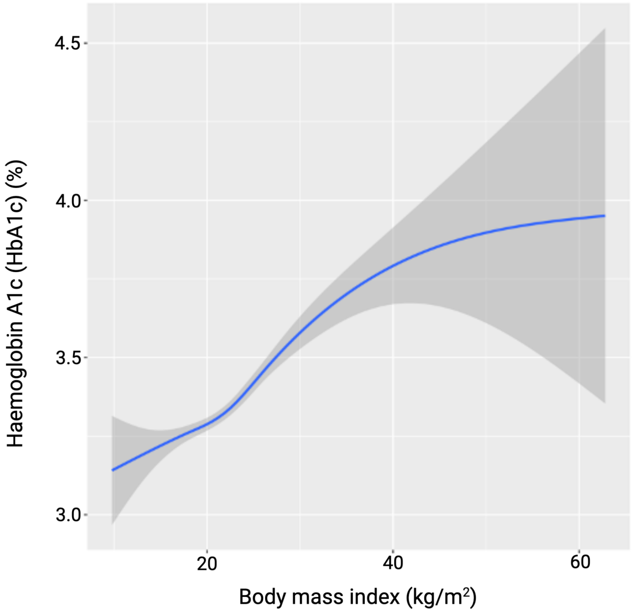
